# Standardized, repeatable ulcerative colitis histology scoring and endpoint assessment using an automated, foundation model-based tool

**DOI:** 10.64898/2026.06.09.26355212

**Authors:** Waleed Tahir, John Shamshoian, John Tauber, Lani K. Clinton, Michael Griffin, Chintan Shah, Geetika Singh, Darren Fahy, Kathleen Sucipto, Jacqueline A. Brosnan-Cashman, Tara A. Altepeter, Sabyasachi Bhattacharya, Wallace Crandall, Chong Duan, Jeremy D. Gale, Vandana Gupta, Hélène Haarmann, Noam Harpaz, Andrea Hooper, Julie Horowitz, Andres Hurtado-Lorenzo, Bader E. Hussaini, Vipul Jairath, Autumn Jones, Ben Kostiuk, F. Stephen Laroux, Trevor Lissoos, Russell B. McBride, Fedaa Najdawi, Anil Nayyar, Mark T. Osterman, Pratik Panchal, Darren Ruane, Paresh Thakker, Simon P. Travis, Laura Wilson, Christina Jayson

## Abstract

In clinical trials for ulcerative colitis (UC), pathologists assess disease severity through standardized histological indices, including the Geboes Score, Robarts Histopathology Index (RHI), and Nancy Histologic Index (NHI). Despite strong associations with clinical outcomes, histologic scoring suffers from inter- and intra-reader variability, and consensus criteria for histologic remission remain uncertain. Through a consortium approach, we developed an artificial intelligence-based measurement (AIM) tool for scoring histology in UC mucosal biopsies (AIM-HI UC). This model, trained on a large dataset of UC biopsies (N=10,230), utilizes additive multiple instance learning models leveraging PLUTO, a pathology foundation model, that predict each of the Geboes subgrades, from which the Geboes grade-level score, RHI, and NHI can be calculated. Evaluation of this model on a standalone verification set including clinical trial specimens established algorithm non-inferiority and/or superiority relative to standard qualified pathologists through comparison of algorithm-consensus and pathologist-consensus agreement metrics (non-inferior if difference >-0.1, superior if difference >0, inclusive of confidence intervals). AIM-HI UC was determined to be non-inferior to pathologists (N=3) for the prediction of all seven Geboes subgrades, grade-level Geboes, RHI, NHI, histologic improvement (GS≤3.1), 2A histologic remission (GS<2A.0), and 2B histologic remission (GS<2B.0). AIM-HI UC was superior to pathologists for several Geboes subgrades (GS 0, GS 1, GS 2B, and GS 5), as well as grade-level Geboes, RHI, and positive percent agreement of 2A histologic remission. The model was shown to be greater than 99% repeatable for all histologic scoring metrics examined. Model-derived scores were shown to strongly correlate with canonical histologic features of inflammation, including the proportion of total epithelium that is inflamed (Spearman r=0.83; p<0.01), the proportion of neutrophils localized within crypt epithelium (Spearman r=0.83, p<0.01), and the amount of mucosal area classified as erosion or ulceration (Spearman r=0.80, p<0.01). Overall, these results suggest that AIM-HI UC has the potential to improve consistency of UC histology interpretation, providing a path toward standardization of UC histology scoring in clinical trials.

## Introduction

Treatment goals for ulcerative colitis (UC) have evolved from symptom control to achieving disease clearance [1]. However, reliable biomarkers to predict outcomes or treatment response remain lacking, the development of which could greatly improve patient care [2].

A complicating factor for assessing histologic response to treatment in UC has been the lack of standardized criteria. Endoscopic remission is assessed using the Mayo endoscopic score (MES), with endoscopic remission being defined as MES=1 or=0 [3]. However, compared to endoscopic remission, histologic remission has been observed to be a stronger predictor of clinical relapse in UC, and residual inflammation (not observable with endoscopic techniques) is associated with increased surgical intervention and colorectal neoplasia [4–6]. Furthermore, histologic remission has been shown to better predict clinical outcomes than endoscopic remission [4,5,7–12]. Neutrophilic inflammation, in particular, has been shown to be predictive of disease activity, including relapse [6,13–17]. However, despite the clear promise of histologic metrics for UC evaluation, a standardized definition of histologic remission remains lacking, even as the United States Food and Drug Administration has recommended the inclusion of histologic remission as a secondary or exploratory endpoint in UC clinical trials [18].

UC histology is scored using several histologic indices, including the Geboes Score [19], Robarts Histopathology Index (RHI) [20], and Nancy Histologic Index (NHI) [21]. When assigning these scores, pathologists assess the underlying composition of the UC inflammatory microenvironment. While recommendations of which scoring system to use vary [21,22], GS, RHI and NHI were found to be similarly reliable when scored by sub-specialty trained GI pathologists and were able to assess histologic changes due to treatment [23]. That said, the manual, visual evaluation of histologic slides by pathologists can be complicated by inter- and intra-reader variability, especially among general pathologists when evaluating cases with less severe histology [20,23]. The reliance on GI-pathology trained pathologists limits the pool of scorers for clinical trials and highlights the opportunity for tools to improve scoring even in routine clinical practice to guide treatment decisions.

Artificial intelligence (AI) models have been increasingly utilized for assessment of digitized histology specimens across pathology, including in UC. Algorithms to predict NHI scores from digitized whole slide images (WSIs) of hematoxylin and eosin (H&E)-stained UC biopsies have been shown to have strong concordance with pathologists [24,25], while approaches to predict tissue regions and cell types of the UC microenvironment from these input images have allowed greater interpretability of these predictions [24]. Recent advances in foundation models – AI systems pre-trained on vast, diverse histopathology image datasets to learn generalizable tissue representations [26,27] – have the potential to yield AI-enabled digital pathology approaches that are highly performant and strongly generalizable for WSI scoring and analysis.

We assembled a cross-industry and academic consortium to build a foundation model-based tool to generate automated histologic scores from H&E-stained biopsies and improve histologic endpoint assessment in UC clinical trials. The goals of this study were to 1) assess the performance of this tool compared to visual pathologist scoring, 2) examine the repeatability of the model, and 3) evaluate the interpretability of model-derived predictions.

## Methods and Materials

### Ethics

This study was conducted in accordance with the Declaration of Helsinki and Good Clinical Practice guidelines. Anonymized digitized whole slide images (WSIs) of H&E-stained intestinal biopsies were obtained from adult patients with UC that had participated in the following complete clinical trials of UC therapeutics: NCT02958865 [28], NCT02840721 [29], NCT02407236 [30], and NCT002209456 [31]. Approval by central institutional review boards was previously described, and all patients had provided informed consent for future research and tissue histology [28–31]. Additional biopsy WSIs were obtained from Mt. Sinai Hospital (New York, New York; IRB# 21-00714) and PathAI Diagnostics (Memphis, Tennessee; IRB# 20214284). The final sets of WSIs used herein were generously provided by Cleveland Clinic Foundation (CCF; Cleveland, Ohio) under a data licensing arrangement approved by the CCF Institutional Review Board, or were obtained from South Bend Medical Foundation (South Bend, Indiana), and Discovery Life Sciences (Huntsville, Alabama).

### Datasets

A total of N=10,230 anonymized, H&E-stained UC biopsies were digitized using the Aperio AT2 (Leica Biosystems, Deer Park, IL), Aperio GT450 (Leica Biosystems, Deer Park, IL), Huron LE 120 (St. Jacobs, ON, Canada), and Hamamatsu NanoZoomer S210 (Hamamatsu Photonics, Bridgewater, NJ) slide scanners. WSIs were split into training (N=7466, 73%), optimization (N=2186, 21.4%) and held-out test (N=578, 5.6%) sets. An additional standalone analytical verification (SAV) dataset was constructed of specimens from two datasets: WSIs (N=533) scanned on AT2 from a data source not used for model training and WSIs (N=451) scanned on GT450 from a data source used in training. Characteristics of these datasets are listed in **Table 1**.

**Table 1.** Patient demographics.

|  |  | **Training &**<br>**optimization**<br>**datasets**<br>**N<sub>WSI</sub>=9,652** | **Held-Out Test set**<br>**N<sub>WSI</sub>=578** | **Standalone**<br>**verification set**<br>**N<sub>WSI</sub>=985** |
| --- | --- | --- | --- | --- |
| Scanner | Aperio AT2 | 53.97% | 23.18% | 54.11% |
|  | Aperio GT450 | 27.78% | 0% | 45.89% |
|  | Huron LE 120 | 10.55% | 46.37% | 0% |
|  | Hamamatsu NanoZoomer S210 | 7.71% | 30.45% | 0% |
| Collection Type | Biopsy | 100% | 100% | 100% |
| Data Source | Clinical Source 1 | 37.51% | 0% | 0% |
|  | Clinical Source 2 | 26.24% | 23.18% | 0% |
|  | Clinical Source 3 | 0.79% | 0% | 0% |
|  | Clinical Source 4 | 16.68% | 0% | 45.89% |
|  | Clinical Source 5 | 10.55% | 46.37% | 0% |
|  | Clinical Source 6 | 7.71% | 30.45% | 0% |
|  | Clinical Source 7 | 0.53% | 0% | 0% |
|  | Clinical Source 8 | 0% | 0% | 54.11% |
| Sex | Male | 30% | 19.9% | 56.8% |
|  | Female | 32% | 15.1% | 43.2% |
|  | Unspecified | 38% | 65% | 0% |
| Geboes grade-level Score | 0 | 2.51% | 30.3% | 18.5% |
|  | 1 | 0.60% | 4.3% | 5.7% |
|  | 2 | 3.02% | 14.9% | 15.4% |
|  | 3 | 3.90% | 10.7% | 14.8% |
|  | 4 | 3.85% | 11.9% | 14.9% |
|  | 5 | 8.63% | 27.9% | 30.7% |
|  | Discordant | 3.60% | 0% | 0% |
|  | Unavailable | 74.26% | 0% | 0% |

### Model Development

AIM-HI UC v2.0* is a model trained to predict histologic scores from H&E-stained UC biopsies, as described below. To train this model, gastrointestinal pathologists specializing in gastrointestinal pathology (N=28 total; N=3 per WSI) provided slide-level Geboes subgrade scores for each WSI in the training set. The median consensus score was used to determine ground-truth Geboes subscores for each slide, which were used to train patch-based multiple instance learning (MIL) classifiers [32] for patch-based classification of each Geboes subgrade (i.e., seven total models to predict GS 0, GS 1, GS 2A, GS 2B, GS 3, GS 4, and GS 5). These classifiers incorporated a backbone leveraging PLUTO, a pathology foundation model [33]. Each MIL model yielded a primary output of scores for each Geboes subgrade. From these values, the Geboes grade-level score could be derived, as could the RHI [20,34] and NHI [21].

These models were trained using an iterative process in which successive models were benchmarked to an optimization set. Briefly, model performance was compared to pathologist scores on this dataset in a nested pairwise manner [35] to assess model non-inferiority. For each predicted output, the model was determined to be non-inferior to a standard qualified pathologist if, for the difference between model and pathologist quadratic kappa values, the lower bound of the 95% confidence interval was greater than -0.1. Harmonized definitions for potentially subjective subgrade scores were established, and additional training sessions were held for scoring pathologists. This cycle was repeated until model performance converged to a level non-inferior to pathologists on the optimization set.

### Model Evaluation

Geboes slide-level and subgrade scores were collected from pathologists (N=29) and compared to model predictions in the SAV set and held-out test set using inter-pathologist pairwise agreement metrics [35] and complementary ML-based pairwise agreement metrics, as follows. Quadratic kappa (Geboes subgrades) and linear kappa (grade-level Geboes, RHI, NHI) were used to assess subgrade predictions and derived grade-level GS, NHI and RHI relative to study pathologists, respectively. Positive percent agreement (PPA; measuring the proportion of positive calls by the model among slides scored positive by the comparator pathologist) and negative percent agreement (NPA; measuring the proportion of negative calls by the model scored negative by the comparator pathologist) were used to assess histologic improvement (GS <u>≤</u> 3.1; less than 5% neutrophil infiltration, no crypt destruction, and no erosions or ulcers), 2A histologic remission (GS<2A.0; no eosinophils or neutrophils in lamina propria), and 2B histologic remission (GS<2B.0; no neutrophils in lamina propria). PPA and NPA were chosen in lieu of sensitivity and specificity metrics given the known inter-pathologist variability in the reference standard.

For each predicted output, AIM-HI UC was determined to be non-inferior to a standard qualified pathologist if the agreement between AIM-HI UC and consensus was sufficiently close to the average agreement among standard qualified pathologists and consensus. Specifically, non-inferiority is achieved when the difference between these two values, inclusive of the entire confidence interval, exceeds -0.1. In addition, AIM-HI UC will be considered superior when the difference between these two values, inclusive of the entire confidence interval, exceeds 0.

### Model Repeatability Analysis

AIM-HI UC was deployed on each slide in the SAV dataset (N=985) across ten independent deployments, and the following metrics were captured: Geboes subgrades, overall grade-level Geboes score, RHI, NHI, histologic improvement, 2A histologic remission, and 2B histologic remission. For each metric, the percentage of slides for which complete agreement was obtained was calculated to determine model repeatability.

### Quantification of UC histology features

IBDExplore v2.0* (PathAI, Boston, MA) is a suite of models that, together, provides comprehensive characterization of the UC microenvironment by excluding regions of model-predicted background and artifact (**Supplementary Table 1)** and subsequently quantifying tissue regions (**Supplementary Table 2**), cell types (**Supplementary Table 3**), goblet cell mucin (**Supplementary Table 4**), crypts (**Supplementary Table 5)**, and crypt subtypes (**Supplementary Table 6**). Tissue regions predicted by the model include epithelium (normal and infiltrated), crypt lumen, crypt abscess, muscularis mucosa, lamina propria, erosion or ulceration, granulation tissue, lymphoid aggregates, goblet cell mucin, and blood vessels. Cell nuclei were exhaustively classified as neutrophils, eosinophils, lymphocytes (intraepithelial and non-intraepithelial), enterocytes, goblet cell nuclei, plasma cells, fibroblasts, or macrophages. Crypts were classified as normal or inflamed.

Human interpretable features (HIFs) quantifying these histologic features were extracted as previously described [34]. In short, features capturing raw and relative counts, densities, areas, and spatial relationships across cell types and tissue regions were generated through the combination of model heatmap overlays. IBDExplore HIFs were compared to median consensus pathologist and AIM-HI UC Geboes subgrade and grade level scores using spearman correlation.

*AIM-HI UC and IBDExplore are For Research Use Only. Not for use in diagnostic procedures.

### Statistical Analysis

All comparisons of AIM-HI UC predictions to pathologist scores were calculated using nested pairwise agreement metrics [35], as follows: Geboes subgrades were assessed using quadratic kappas, while Geboes overall grade-level scores, RHI, and NHI were assessed using linear kappas. The consensus for each WSI was defined as the median pathologist score. Accuracy among pathologists and AIM-HI UC in discerning histologic remission and improvement criteria was quantified as positive percent agreement (PPA) and negative percent agreement (NPA). All confidence intervals were generated by bootstrapping with 2000 iterations. IBDExplore HIFs were compared to median consensus pathologist and AIM-HI UC Geboes subgrade and grade level scores using spearman correlation. All analyses were completed in the Python 3.10 programming language, using scipy, sklearn, pandas, and numpy packages.

## Results

### Accuracy of AI-derived UC histology scoring

We developed AIM-HI UC to directly predict Geboes subgrade scores, from which Geboes grade-level scores, RHI, and NHI can be calculated (**Figure 1**). Histologic improvement, 2A histologic remission, and 2B histologic remission can be calculated from AIM-HI UC outputs, as well. To assess the accuracy of the model, we compared model outputs to pathologist-derived scores.

**Figure 1.**
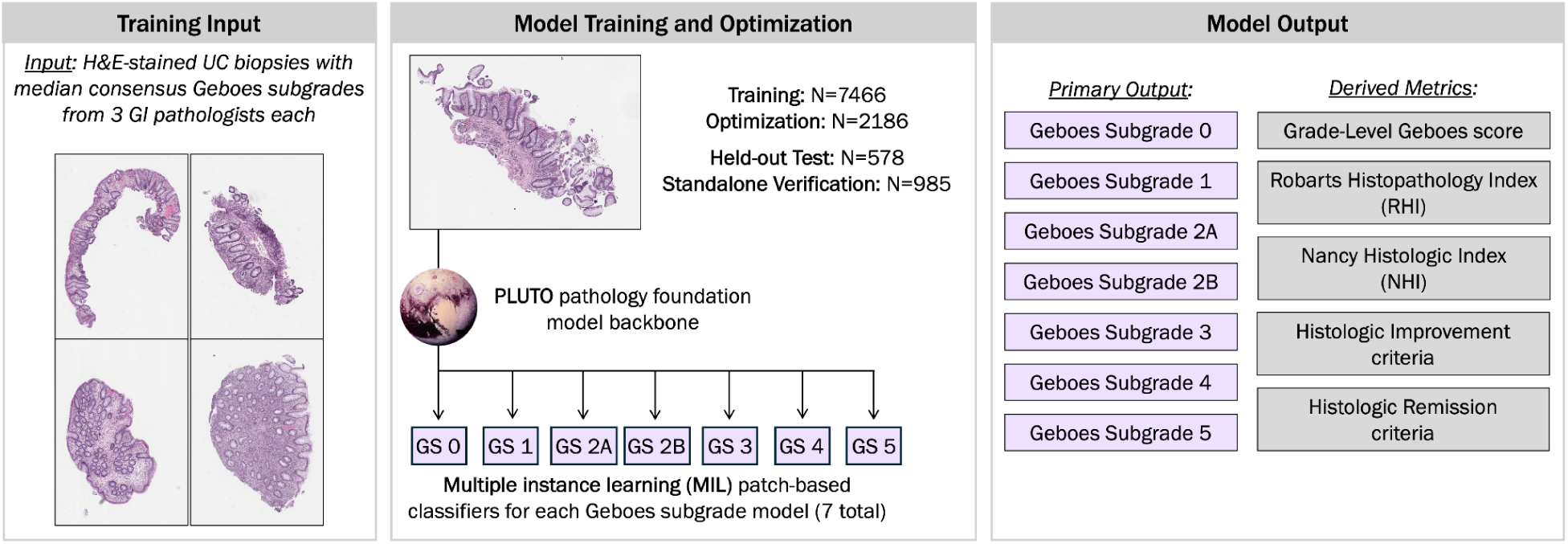
AIM-HI UC training process and output.

#### Accuracy in held out standalone analytical verification dataset

Model-predicted and pathologist-derived Geboes subgrade scores were compared in the overall SAV dataset (**Table 2**; **Figure 2A-B**). As shown in **Figure 2B** and **Table 2**, the model met non-inferiority criteria (the lower bound of the 95% confidence interval was greater than -0.1 for the difference between model and pathologist kappa values) for all Geboes subgrades, while it achieved superior performance (the lower bound of the 95% confidence interval was greater than 0.0 for the difference between model and pathologist kappa values) to pathologists for GS0, GS1, GS2B, and GS5. GS2A is the only subgrade where the quadratic kappa for the model was less than that of pathologists, indicating that pathologists agreed with each other more than the model agreed with pathologists, while both the model and pathologist quadratic kappa was lowest for GS4. Inter-rater variability is known to be particularly prevalent for pathologist scoring of GS2A and GS4, which may have contributed to these results [20,36].

**Figure 2.**
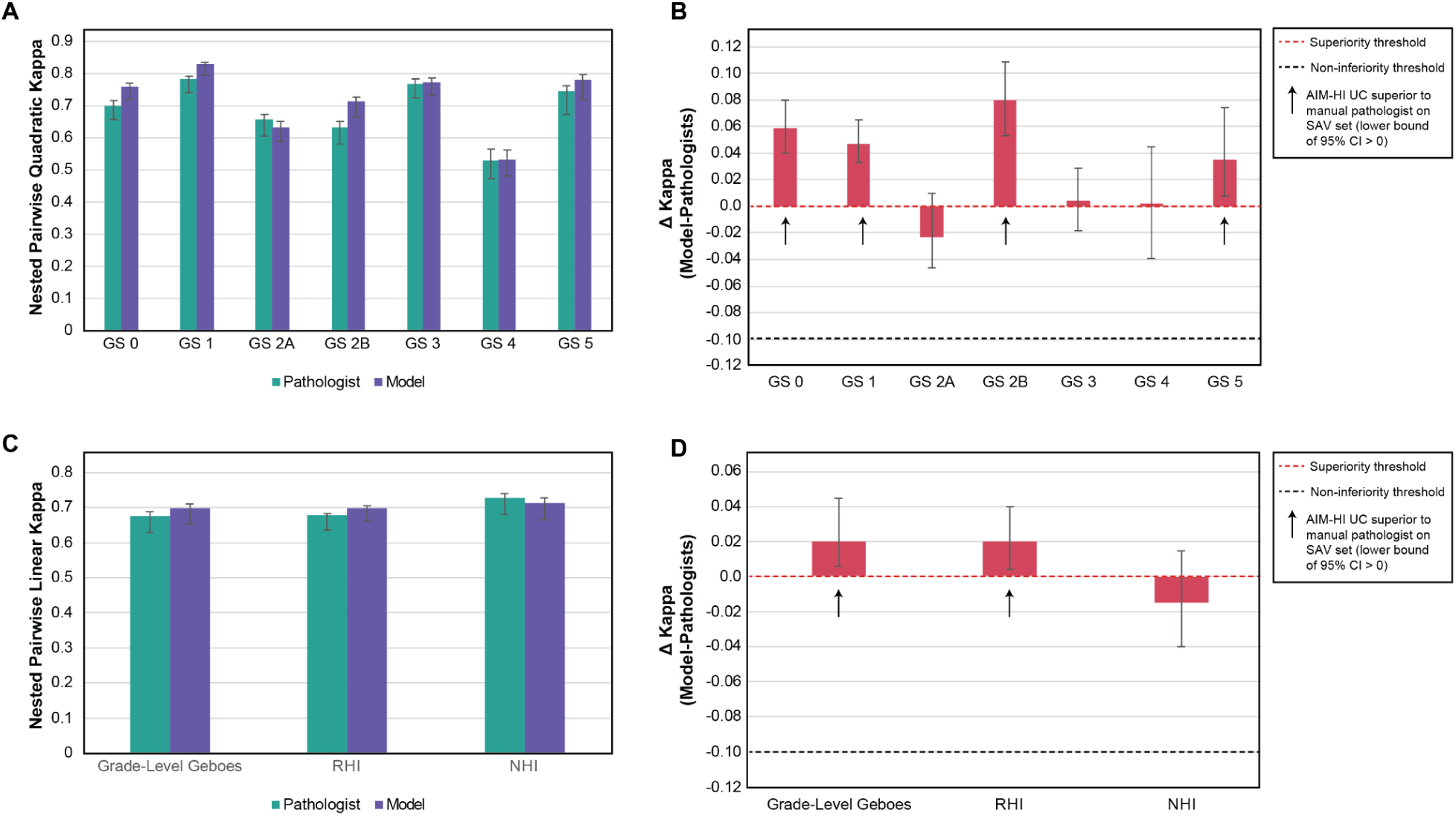
AIM-HI UC model performance compared to pathologists for histologic scoring. A) Comparison of nested pairwise quadratic kappa values from pathologist manual scoring and AIM-HI UC predictions of Geboes subgrades. B) Difference in quadratic kappa between model and pathologist for Geboes subgrade determination. C) Comparison of nested pairwise linear kappa values from pathologist manual scoring and AIM-HI UC predictions of overall grade-level Geboes score, RHI, and NHI. D) Difference in linear kappa between model and pathologist for overall grade-level Geboes score, RHI, and NHI determination. Error bars reflect 95% confidence intervals. GS: Geboes score, RHI: Robarts Histopathology Index, NHI: Nancy Histology Index.

**Table 2.** Comparison of manual pathologist Geboes subgrade scores with model predictions in the full SAV dataset.

| **Score** | **Pairwise pathologist QK**<br>**(95% CI)** | **Pairwise model QK**<br>**(95% CI)** | **Difference**<br>**(95% CI)** |
| --- | --- | --- | --- |
| GS 0 | 0.701<br>(0.656, 0.715) | 0.760<br>(0.719, 0.770) | 0.059<br>(0.040, 0.080) |
| GS 1 | 0.783<br>(0.741, 0.792) | 0.830<br>(0.795, 0.836) | 0.047<br>(0.033, 0.065) |
| GS 2A | 0.656<br>(0.604, 0.672) | 0.633<br>(0.589, 0.652) | -0.023<br>(-0.046, 0.010) |
| GS 2B | 0.632<br>(0.580, 0.652) | 0.712<br>(0.665, 0.728) | 0.08<br>(0.053, 0.109) |
| GS 3 | 0.768<br>(0.723, 0.784) | 0.773<br>(0.732, 0.786) | 0.004<br>(-0.018, 0.029) |
| GS 4 | 0.530<br>(0.474, 0.565) | 0.532<br>(0.481, 0.563) | 0.002<br>(-0.039, 0.045) |
| GS 5 | 0.747<br>(0.672, 0.762) | 0.782<br>(0.719, 0.798) | 0.035<br>(0.008, 0.074) |

QK: quadratic kappa; GS: Geboes Score; CI: Confidence Interval

Model-derived calculations of grade-level Geboes score, NHI, and RHI, were also compared to pathologist scores (**Table 3**; **Figure 2C-D**). Here, as shown in **Figure 2D** and **Table 3**, the model met non-inferiority criteria for grade-level Geboes, RHI, and NHI and achieved superior performance to pathologists for grade-level Geboes and RHI (**Figure 2D**). Notably, the linear kappa for the model was greater than that of pathologists for grade-level Geboes and RHI, while it was slightly less for NHI.

**Table 3.** Comparison of manual pathologist grade-level Geboes scores, RHI, and NHI with model predictions in the full SAV dataset.

| **Score** | **Pairwise pathologist LK**<br>**(95% CI)** | **Pairwise model LK**<br>**(95% CI)** | **Difference**<br>**(95% CI)** |
| --- | --- | --- | --- |
| Grade-level<br>GS | 0.677<br>(0.629, 0.688) | 0.697<br>(0.653, 0.711) | 0.020<br>(0.006, 0.045) |
| RHI | 0.678<br>(0.637, 0.684) | 0.698<br>(0.660, 0.705) | 0.020<br>(0.004, 0.040) |
| NHI | 0.727<br>(0.680, 0.740) | 0.712<br>(0.666, 0.729) | -0.015<br>(-0.040, 0.015) |

LK: Linear kappa; RHI: Roberts Histopathology Index; NHI: Nancy Histologic Index; CI: Confidence Interval

We also assessed the PPA and NPA of the model for scoring histologic improvement (GS≤3.1), 2A histologic remission (GS<2A.0), and 2B histologic remission (GS<2B.0) in the SAV set and compared these accuracy metrics to manual, visual scoring by pathologists. As shown in **Table 4** and **Figure 3A**, the model achieved superior PPA for measurement of 2A histologic remission compared to pathologists while NPA metrics were non-inferior to pathologists. For 2B histologic remission, the model achieved non-inferior PPA to pathologists. NPA was also non-inferior, although a slight decrease was observed for the model based on the point estimate (**Table 4**, **Figure 3B**). Finally, pathologist PPA was slightly higher than model PPA for measuring histologic improvement, while NPA values were similar (**Table 4**, **Figure 3C**). Thus, the model was deemed non-inferior to pathologists for the prediction of histologic improvement, 2A histologic remission, and 2B histologic remission and achieved superior PPA than pathologists for 2A histologic remission.

**Figure 3.**
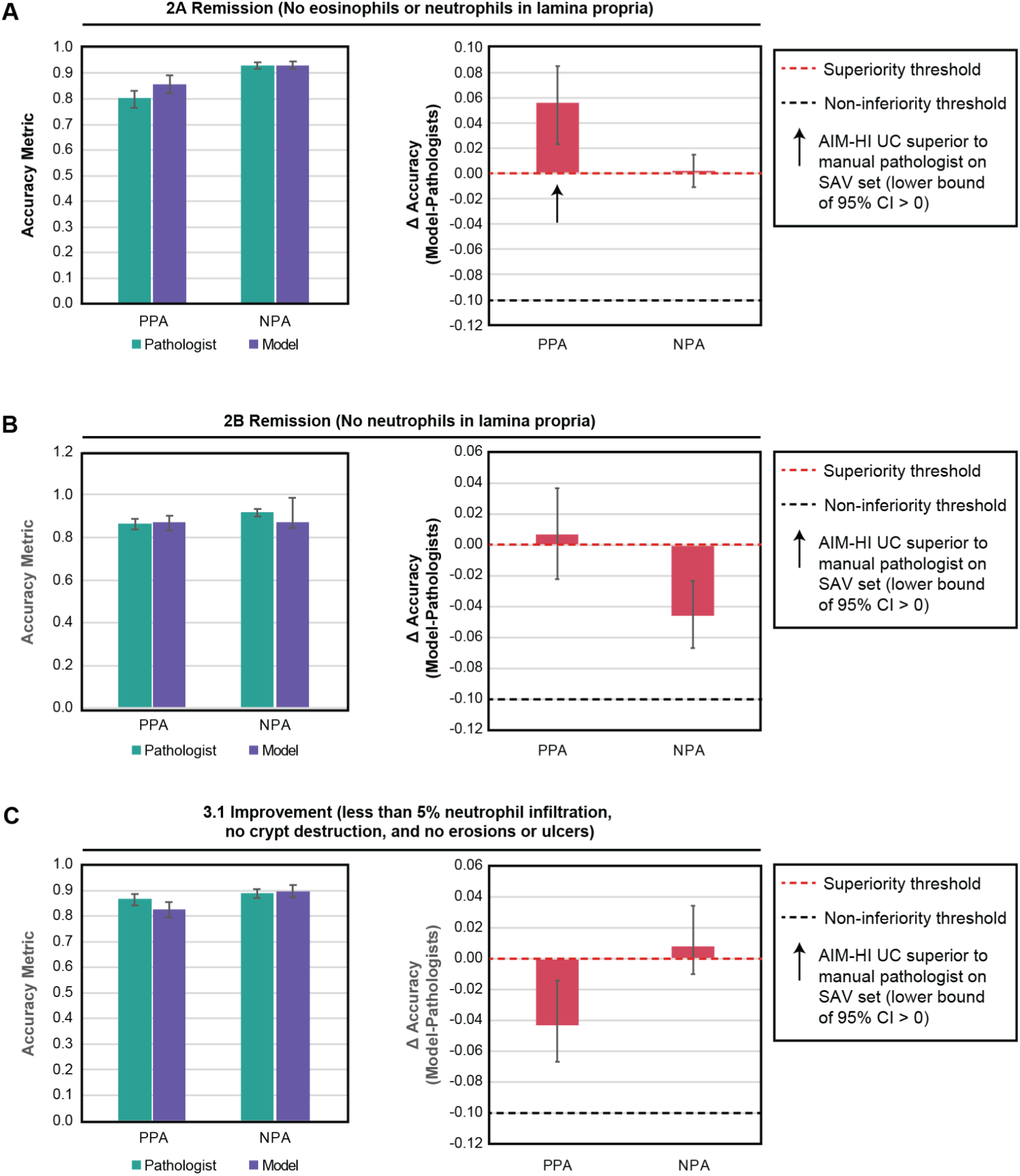
AIM-HI UC model performance compared to pathologists for determination of histologic remission or improvement status. A) Comparison of positive percent agreement (PPA) and negative percent agreement (NPA) from pathologist manual scoring and AIM-HI UC prediction of 2A histologic remission. Difference in PPA and NPA between model and pathologist for 2A histologic remission is also shown. B) Comparison of PPA and NPA from pathologist manual scoring and AIM-HI UC prediction of 2B histologic remission. Difference in PPA and NPA between model and pathologist for 2B histologic remission is also shown. C) Comparison of PPA and NPA from pathologist manual scoring and AIM-HI UC prediction of histologic improvement. Difference in PPA and NPA between model and pathologist for histologic improvement is also shown. Error bars reflect 95% confidence intervals. PPA: positive percent agreement, NPA: negative percent agreement; CI: confidence interval.

**Table 4.** Comparison of manual pathologist assessment of histologic remission and improvement with model predictions in the full SAV dataset.

| **Metric** |  | **Pairwise pathologist**<br>**(95% CI)** | **Pairwise model**<br>**(95% CI)** | **Difference**<br>**(95% CI)** |
| --- | --- | --- | --- | --- |
| 2A Histologic Remission | PPA | 0.802<br>(0.765, 0.832) | 0.856<br>(0.822, 0.893) | 0.056<br>(0.027, 0.089) |
|  | NPA | 0.930<br>(0.916, 0.943) | 0.931<br>(0.916, 0.947) | 0.002<br>(-0.011, 0.015) |
| 2B Histologic Remission | PPA | 0.864<br>(0.838, 0.887) | 0.871<br>(0.836, 0.904) | 0.007<br>(-0.022, 0.037) |
|  | NPA | 0.919<br>(0.900, 0.932) | 0.874<br>(0.846, 0.896) | -0.046<br>(-0.067, -0.023) |
| Histologic Improvement | PPA | 0.867<br>(0.843, 0.885) | 0.825<br>(0.794, 0.854) | -0.043<br>(-0.067, -0.014) |
|  | NPA | 0.889<br>(0.870, 0.904) | 0.897<br>(0.875, 0.921) | 0.008<br>(-0.01, 0.034) |

PPA: positive percent agreement; NPA: negative percent agreement; CI: confidence interval

#### Scanner-specific accuracy

To more comprehensively assess model generalizability across scanners, we compared histology scores manually provided by pathologists to model-derived scores on slides from the standalone analytical verification set, which were digitized using the GT450 or AT2 slide scanners. For both the GT450 and AT2, the model was deemed non-inferior to pathologists for the prediction of all seven Geboes subgrades (**Supplementary Table 7; Supplementary Figure 1A-B**), as well as grade-level Geboes, RHI, and NHI (**Supplementary Table 8; Supplementary Figure 1C-D**).

Similar scanner-specific evaluations were also performed for histologic remission and improvement criteria. Here, again, for both the GT450 and AT2, the model was deemed non-inferior to pathologists for the prediction of 2A histologic remission (**Supplementary Table 9; Supplementary Figure 2A-B**, 2B histologic remission (**Supplementary Table 9; Supplementary Figure 2C-D**, and histologic improvement (**Supplementary Table 9; Supplementary Figure 2E-F**).

#### Accuracy in a held-out dataset

To further characterize model performance, we compared the outputs to pathologist scores in a separate held-out test set (N=578) composed of specimens from data sources used in training and imaged using multiple scanners (AT2, Huron, Hamamatsu). In this dataset, the model was considered non-inferior to pathologists for all Geboes subgrades, grade-level Geboes, RHI, and NHI, as well as 2A histologic remission, 2B histologic remission, and histologic improvement (**Supplementary Figure 3**). Model performance was superior to pathologists for Geboes subgrades 0, 1, 2A, 2B, 3, and 5, as well as grade-level Geboes, RHI, NHI,

Collectively, the model was determined to be non-inferior to pathologists for all UC scoring parameters assessed across multiple scanners and two held-out datasets. These results support the robustness and broad applicability of the model for histologic scoring in future clinical trial settings.

### Repeatability of AI-derived UC histology scoring

A critical potential advantage of AI-based histology scoring is the elimination of intra-reader variability. To assess the repeatability of AIM-HI UC, each WSI from the SAV dataset (N=985) was evaluated by the model ten times. The degree to which scores were in agreement were assessed, and results are shown in **Table 5** and **Figure 4**. For all collected metrics, AIM-HI UC had a greater than 99% repeatability across ten independent model deployments. These repeatability values are in contrast to previously reported intra-pathologist agreement for Geboes subgrades and grade-level scores, RHI, and NHI ranging from 72-94% [20]. These results demonstrate the ability of AIM-HI UC to provide greater consistency to UC histologic scoring.

**Figure 4.**
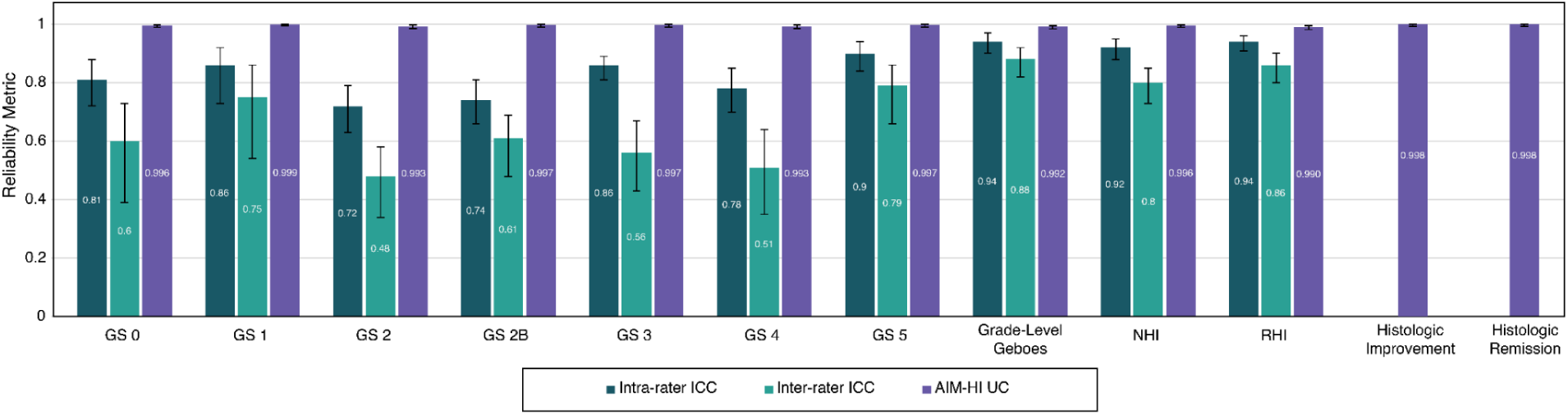
Repeatability of AIM-HI UC outputs compared to historical manual data. AIM-HI UC repeatability was assessed by deploying the algorithm ten times on the same whole slide images. Historical intra-rater and inter-rater ICC values were obtained for Geboes subgrades,^17^ grade-level Geboes,^19^ RHI,^19^ and NHI.^19^ Error bars represent 95% confidence intervals.

**Table 5.** Algorithm repeatability assessment using 10 algorithm deployments per WSI.

| **Metric** | **Total WSIs** | **WSIs in complete agreement** | **Repeatability (95% CI)** |
| --- | --- | --- | --- |
| GS 0 | 985 | 981 | 0.996 (0.9896, 0.9984) |
| GS 1 | 985 | 984 | 0.999 (0.9943, 0.9998) |
| GS 2A | 985 | 978 | 0.993 (0.9854, 0.9966) |
| GS 2B | 985 | 982 | 0.997 (0.9911, 0.999) |
| GS 3 | 985 | 982 | 0.997 (0.9911, 0.999) |
| GS 4 | 985 | 978 | 0.993 (0.9854, 0.9966) |
| GS 5 | 985 | 982 | 0.997 (0.9911, 0.999) |
| Grade-level GS | 985 | 977 | 0.992 (0.9841, 0.9959) |
| NHI | 985 | 981 | 0.996 (0.9896, 0.9984) |
| RHI | 985 | 975 | 0.99 (0.9814, 0.9945) |
| Histologic Improvement | 985 | 983 | 0.998 (0.9926, 0.9994) |
| 2A Histologic Remission | 985 | 983 | 0.998 (0.9926, 0.9994) |
| 2B Histologic Remission | 985 | 985 | 1.0 (0.9961, 1.0) |

### Interpretability of AI-derived UC histology scoring

To validate that AIM-HI UC predictions capture biologically relevant histologic features, we assessed the correlation between model-derived scoring predictions and quantitative AI-derived features of the UC inflammatory microenvironment in the held-out test set, predicted using IBDExplore (**Figure 5A**). We first assessed the correlation between AI-derived Geboes subgrades and selected AI histology features meant to capture each (**Supplemental Table 10**). As expected, these features were significantly associated with increasing Geboes subscores (**Supplemental Figure 4**), supporting the interpretability of model predictions. We next assessed features capturing canonical inflammation across model-predicted Geboes grade-level scores. As expected, we observed strong positive correlations between AI-predicted Geboes grade-level scores and these features, including the relative area of total epithelium that is inflamed (**Figure 5B**, Spearman r=0.83; p<0.01) and the proportion of total neutrophils localized within crypt epithelium (**Figure 5C**, Spearman r=0.83, p<0.01), and the relative mucosal area classified as erosion or ulceration (**Figure 5D**, Spearman r=0.80, p<0.01). In contrast, we noted inverse correlations between the relative mucosal area classified as normal epithelium and Geboes grade-level score (**Figure 5E**, Spearman r=-0.62, p<0.01), Together, these trends support the notion that canonical inflammation accompanies the histologic progression of UC, as captured by AIM-HI UC-generated grade-level Geboes scores.

**Figure 5.**
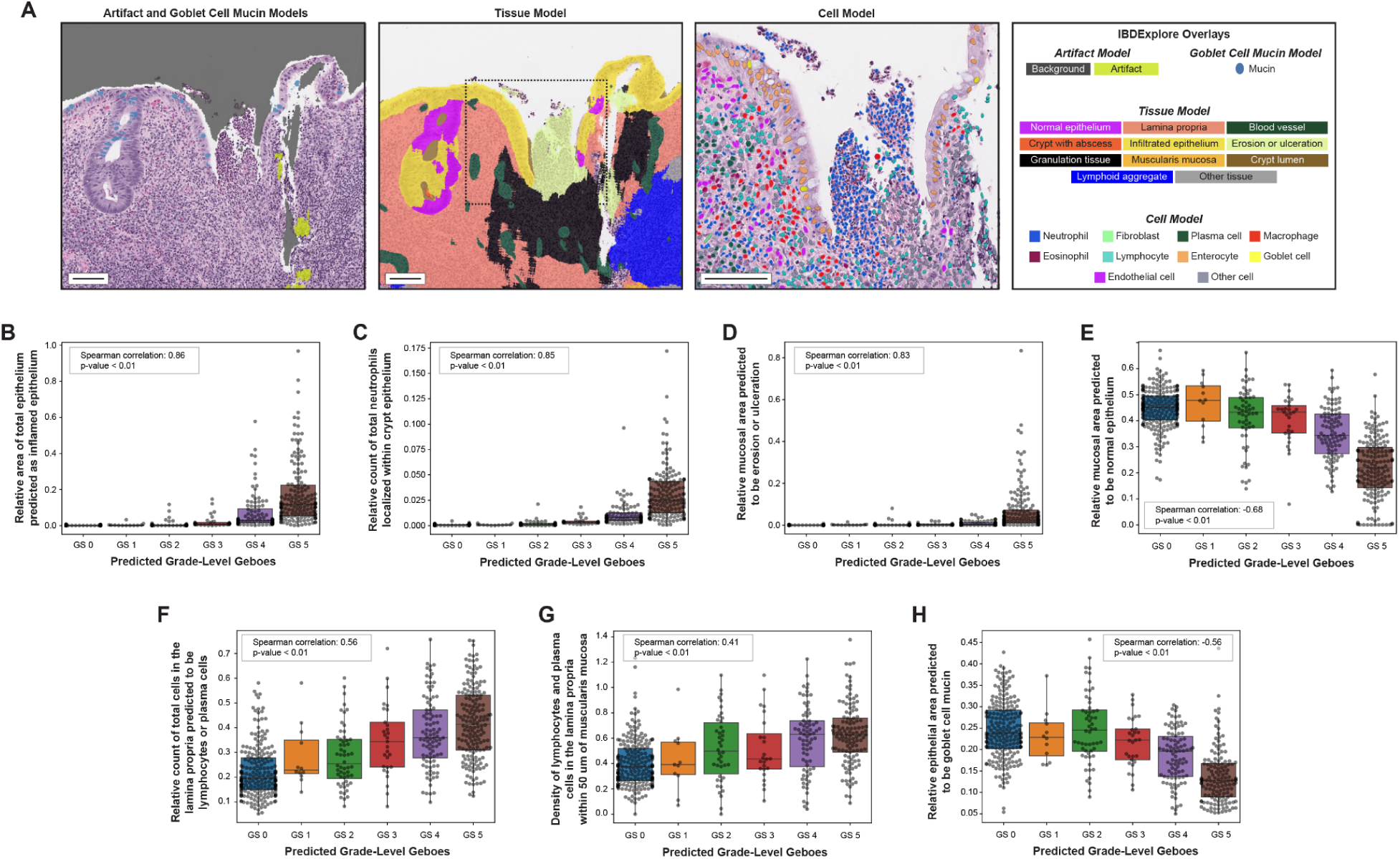
Correlation between AIM-HI UC scores and AI-quantified features of the inflammatory microenvironment. A) Schematic of features quantified by IBDExplore. Scale bars represent 100 μm. B-F) Correlation between AIM-HI UC-predicted Geboes grade-level scores and the following IBDExplore-derived histology features: B) relative area of total epithelium predicted as inflamed epithelium, C) relative count of total neutrophils localized within crypt epithelium, D) relative mucosal area predicted to be erosion or ulceration, E) relative mucosal area predicted to be normal epithelium, F) relative count of lymphocytes and plasma cells localized within the lamina propria, G) density of lymphocytes and plasma cells in the lamina propria within 50 μm of the muscularis mucosa, and H) relative area of total epithelium predicted to be goblet cell mucin. Error bars reflect 95% confidence intervals. GS: Geboes Score.

UC histology is not completely characterized by the Geboes scoring criteria. To better understand how additional histology features capturing distinct aspects of UC biology relate to Geboes score, we assessed the degree of association between these features and model-predicted scores. First, we assessed the association of features indicative of chronic inflammatory infiltrate with model-predicted Geboes grade-level scores. The proportion of total cells in the lamina propria predicted to be lymphocytes, eosinophils, and plasma cells was positively correlated with increasing Geboes grade-level score (**Figure 5F**; Spearman r=0.57, p<0.01). When examined individually, weaker degrees of correlation with grade-level Geboes were observed for lymphocytes (**Supplemental Figure 5A;** Spearman r=0.45, p<0.01), eosinophils (**Supplemental Figure 5B;** Spearman r=0.27, p<0.01), and plasma cells (**Supplemental Figure 5C;** Spearman r=0.47, p<0.01). Next, we assessed the association of basal lymphoplasmacytosis, a known hallmark of chronic inflammation in UC [37], with Geboes grade-level scores. To make this comparison, we used an IBDExplore feature with a predefined spatial distance threshold, the density of lymphocytes and plasma cells in lamina propria localized within 50 μm of muscularis mucosa, to represent the anatomical changes that occur in basal lymphoplasmacytosis. We observed a significant positive correlation of this feature with increasing Geboes grade-level scores, as well (**Figure 5G**; Spearman r=0.56, p<0.01). Together, these results support the known phenomenon of chronic inflammation occurring with increasing UC severity. Lastly, given the known protective function of colonic mucin for epithelial barrier health [38], we assessed the association of model-predicted goblet cell mucin with increasing Geboes grade-level scores. As shown in **Figure 5H**, the area of goblet cell mucin relative to total epithelial area was significantly inversely correlated with model-predicted grade-level Geboes scores (Spearman r=-0.56, p<0.01). These results support the notion that mucin reduction accompanies intestinal barrier dysfunction in progressing UC.

## Discussion

The association between histologic remission and outcomes in UC ^[4,5,7–12]^ highlights the necessity of accurate histologic assessment. However, the manual evaluation of histologic slides by pathologists can be complicated by inter- and intra-reader variability [20,23]. To address these issues, we developed an AI histology tool, AIM-HI UC, to standardize histologic scoring of routine UC biopsies across the major scoring systems and remission/improvement criteria.

The AIM-HI UC model architecture leverages PLUTO, a pathology foundation model [33]. Pathology foundation models have been trained on large, diverse datasets of whole slide images, potentially improving both model generalizability and accuracy of predictions [26,39]. The improved generalizability of pathology foundation models is thought to result from the increased size and diversity of model training sets [27]. Direct comparison of pathology foundation models to earlier-generation model architectures for ovarian cancer subtyping revealed higher performance by the foundation models [40]. While foundation models have been shown to accurately detect the presence of inflammatory bowel diseases [41], to our knowledge, this study is the first to apply a pathology foundation model to a histologic scoring task in UC.

AIM-HI UC performed well relative to manual pathologist scoring in two datasets. Model performance was determined to be non-inferior for all Geboes subgrades, Geboes grade-level scores, histologic improvement, 2A histologic remission, and 2B histologic remission. Furthermore, across the entire SAV dataset, the model performed in a manner superior to pathologists for GS 0, GS 1, GS 2B, GS 5, grade-level Geboes, and RHI. The superior performance of the model may reflect an improved ability of the model to detect histologic changes that pathologists may miss in a visual assessment, such as alterations in goblet cell mucin levels or the presence of rare neutrophils within the epithelium. Furthermore, the model was shown to be greater than 99% repeatable for the predictions of Geboes subgrades, Geboes grade-level scores, histologic improvement, 2A histologic remission, and 2B histologic remission. Previous studies have shown that intra-pathologist reliability varies based on the metric in question, with reliability for Geboes subgrade scores ranging from 72-90%, 92% for grade-level Geboes, 94% for RHI, and 92% for NHI [20,23]

For measurements of histologic improvement and remission, we elected to use positive, and negative percent agreement (PPA and NPA) rather than sensitivity and specificity. This decision was made given the imperfect reference standard of manual pathologist scoring due to the known inter-pathologist variability. That said, PPA is a metric assessing how well the model identifies true positives and NPA is a metric assessing how well the model identifies true negatives; therefore, while these metrics are distinct from sensitivity and specificity, they can be interpreted in a similar way. Our results indicate that AIM-HI UC was non-inferior to pathologists for identifying true positives and true negatives for each criterion. In other words, the model is able to identify those in remission or having improved as well as a pathologist can.

Taken together, the improved repeatability across all components and superior performance of AIM-HI UC compared to conventional, manually-provided pathologist scoring across many of the components assessed has the potential to standardize clinical trial histologic scoring in UC. The improved ability to reliably generate histologic scores in UC addresses a clear need in the field, given the importance of histologic remission for disease outcomes and the recommendation by the United States Food and Drug Administration to include histologic response as a secondary or exploratory endpoint in UC clinical trials.

Importantly, the potential integration of AIM-HI UC with complementary digital pathology models provides interpretability to the model outputs. Here, we demonstrate that quantitative histologic features from IBDExplore – a suite of models that characterize the UC inflammatory microenvironment — significantly correlate with corresponding Geboes subgrades, as expected. For example, the relative fractions of total eosinophils and total neutrophils localized in the lamina propria is significantly associated with increasing subgrade scores for GS 2A and GS 2B, respectively. In addition, we demonstrate that features associated with epithelial inflammation were significantly correlated with increasing grade-level Geboes scores. These features included the proportion of total epithelium that was inflamed, the proportion of neutrophils localized within crypt epithelium, and the amount of mucosal area classified as erosion or ulceration. Notably, previous work had demonstrated the association between pathologist-derived Geboes grade-level and subgrade scores with AI-derived features of the UC with pathologist-derived scoring to be similar to the observed association with AI-derived scores [42].

Features associated with chronic inflammation (the proportion of lymphocytes, eosinophils, and plasma cells in the lamina propria) and representative of possible basal lymphoplasmacytosis (the density of lymphocytes and plasma cells in lamina propria within 50 μm of muscularis mucosa) were also observed to increase with grade-level Geboes, despite the latter not assessed in the scoring system. Furthermore, features quantifying normal crypt epithelium and goblet cell mucin were inversely correlated with increasing grade-level Geboes scores. The reduction in goblet cell mucin - whose role is typically to coat the epithelium with a mucus barrier [38,43]- with increasing grade-level Geboes scores supports the concept of worsening epithelial barrier health with increasing disease severity. The ability to integrate two tools to measure the amount of goblet cell mucin in parallel with automated Geboes (or Nancy or Robarts) scoring has the potential to inform assessment of mucosal healing, a sought-after treatment target for UC that moves beyond the inflammation to restoration of intestinal lining homeostasis.

This study has several notable strengths, including 1) the foundation model-based digital pathology models used to build the study models, 2) the ability to generate training and testing datasets across large, consortium-driven cohorts, and 3) a large dataset with a unique breadth, representing a spectrum of patients across disease severity. Still, limitations remain. Notably, while we assessed the repeatability of AIM-HI UC in this study, we were not able to evaluate model reproducibility. Future studies will be designed to examine the reproducibility of AIM-HI UC using slides from the same biopsies prepared in different laboratories using distinct scanners, slide staining apparati, etc. Relatedly, additional work is necessary to better understand how the tool performs on multiple biopsies from the same patient, including sequential biopsies and paired biopsies from before and after treatment. Furthermore, some of the features that we identified as being associated with increasing disease severity are sensitive to tissue orientation – most notably, basal lymphoplasmacytosis. The spatial nature of this feature (density of lymphocytes and plasma cells in proximity to muscularis mucosa) renders its measurement dependent on tissue orientation and sectioning angle. Additional evaluation of this feature is needed in independent cohorts of UC. In addition, information around race and ethnicity of patients whose biopsies were included in the datasets was unavailable, limiting our understanding of any potential demographic bias in the model. A recent study revealed that AI pathology models for cancer diagnosis show bias relating to race, gender, and age in nearly one-third of cases [44], highlighting the need for better representation of minority groups in model training. Finally, the dataset used herein was not built to discern treatment effects. As such, future studies will examine how AIM-HI UC compares to manual pathologist scoring in retrospective analyses of clinical trials of therapeutics with different mechanisms of action to discern how the model performs for the detection of treatment-induced changes in histology. In addition, future prospective clinical assessment of the tool is necessary to fully understand how the tool may be incorporated into clinical trial workflows in advance of potential use for assessment of non-exploratory clinical trial endpoints in UC.

Finally, the results presented demonstrate the strong association between AIM-HI UC outputs and conventional pathologist scores. However, we envision that this tool will be incorporated in a manner where pathologists are assisted by this AI tool, rather than the tool being used in isolation. As such, a broader assessment of these “AI-Assisted” reads compared to unassisted pathologists is necessary to determine how pathologists interact with AIM-HI UC, as well as how model assistance improves intra-reader variability and/or allows a single pathologist to perform in line with consensus read.

## Data Availability

Access to feature tables, cell- and tissue- heatmaps will be considered on a case by case basis at the discretion of trial sponsors. Access requests can be made to.
Model parameters for AIM-HI UC and IBDExplore model training, inference, and feature extractions are not disclosed. Access requests for such a code will not be considered to safeguard PathAI intellectual property.

## Acknowledgements

We would like to thank the members of PathAI Pathology Network who contributed towards this work, as well as the software engineering and machine learning operations teams at PathAI for developing the systems and pipelines used for model development. We also acknowledge Anjli Kukreja and Sudha Visvanathan for their contributions to this project. This work was funded by the Foundation for the National Institutes of Health.

## Supplementary Tables

**Supplementary Table 1.** IBDExplore artifact model performance in the held-out test set.

|  | **Pathologist vs.**<br>**Pathologist F1 (95% CI)** | **Model vs. Pathologist**<br>**F1 (95% CI)** | **ΔF1 (95% CI)** |
| --- | --- | --- | --- |
| Evaluable<br>Tissue | 0.92 (0.91, 0.94) | 0.91 (0.89, 0.92) | -0.02 (-0.03, 0) |
| Background | 0.93 (0.91, 0.94) | 0.92 (0.9, 0.94) | -0.01 (-0.02, 0) |
| Artifact | 0.54 (0.42, 0.63) | 0.44 (0.32, 0.53) | -0.09 (-0.19, -0.01) |

Non-inferiority criteria were set based on inter-pathologist variability for each substance which were then evaluated on 375 μm x 375 μm frames (n = 200 frames). Evaluable Tissue and Background performance were considered non-inferior to pathologists if the lower bound of the $\Delta$ F1 confidence interval was $>-0.1$ . Artifact performance was considered non-inferior to pathologists if the lower bound of the $\Delta$ F1 confidence interval was $>-0.2$ .

**Supplementary Table 2.** IBDExplore tissue model performance in the held-out test set.

|  | **Pathologist vs.**<br>**Pathologist F1 (95% CI)** | **Model vs. Pathologist**<br>**F1 (95% CI)** | **$\Delta F1$ (95% CI)** |
| --- | --- | --- | --- |
| Other Tissue | 0.54 (0.32, 0.64) | 0.33 (0.2, 0.42) | -0.21 (-0.28, -0.08) |
| Normal Epithelium | 0.86 (0.85, 0.87) | 0.87 (0.86, 0.88) | 0.01 (0.01, 0.02) |
| Muscularis Mucosa | 0.74 (0.7, 0.78) | 0.74 (0.7, 0.78) | 0 (-0.03, 0.02) |
| Lymphoid Aggregate | 0.64 (0.49, 0.73) | 0.54 (0.37, 0.64) | -0.1 (-0.22, -0.02) |
| Lamina Propria | 0.84 (0.83, 0.86) | 0.84 (0.82, 0.85) | 0 (-0.01, 0) |
| Infiltrated Epithelium | 0.53 (0.49, 0.57) | 0.54 (0.49, 0.57) | 0 (-0.02, 0.02) |
| Granulation Tissue | 0.28 (0.16, 0.38) | 0.35 (0.23, 0.43) | 0.06 (0, 0.11) |
| Erosion or Ulceration | 0.64 (0.44, 0.74) | 0.57 (0.39, 0.67) | -0.07 (-0.13, 0) |
| Crypt Abscess | 0.62 (0.5, 0.7) | 0.58 (0.48, 0.63) | -0.05 (-0.1, 0.01) |
| Crypt Lumen | 0.66 (0.63, 0.69) | 0.65 (0.62, 0.67) | -0.02 (-0.05, 0.01) |
| Blood Vessel | 0.55 (0.52, 0.57) | 0.53 (0.5, 0.55) | -0.02 (-0.04, 0.01) |

Non-inferiority criteria were set based on inter-pathologist variability for each substance which were then evaluated on 375 $\mu\text{m}$ x 375 $\mu\text{m}$ frames (n = 400 frames). Normal epithelium, infiltrated epithelium, crypt lumen, lamina propria, and blood vessel performance were considered non-inferior to pathologists if the lower bound of the $\Delta F1$ confidence interval was $>-0.1$ . Crypt abscess, erosion or ulceration, granulation tissue and other tissue performance were considered non-inferior to pathologists if the lower bound of the $\Delta F1$ confidence interval was $>-0.2$ . Lymphoid aggregate performance was evaluated, but considered an exploratory substance.

**Supplementary Table 3.** IBDExplore cell model performance in the held-out test set.

|  | **Pathologist vs.**<br>**Pathologist F1 (95% CI)** | **Model vs. Pathologist**<br>**F1 (95% CI)** | **ΔF1 (95% CI)** |
| --- | --- | --- | --- |
| Plasma Cells | 0.55 (0.52, 0.58) | 0.59 (0.56, 0.62) | 0.04 (0.03, 0.05) |
| Other Cells | 0.28 (0.26, 0.3) | 0.3 (0.28, 0.32) | 0.02 (0, 0.04) |
| Neutrophils | 0.5 (0.45, 0.54) | 0.51 (0.46, 0.55) | 0.01 (-0.02, 0.03) |
| Macrophages | 0.1 (0.07, 0.13) | 0.11 (0.09, 0.14) | 0.02 (-0.01, 0.03) |
| Lymphocytes | 0.52 (0.5, 0.54) | 0.52 (0.5, 0.54) | 0 (-0.01, 0.03) |
| Goblet Cells | 0.4 (0.36, 0.43) | 0.42 (0.38, 0.45) | 0.02 (0, 0.05) |
| Fibroblasts | 0.38 (0.34, 0.41) | 0.38 (0.35, 0.41) | 0 (-0.02, 0.03) |
| Eosinophils | 0.61 (0.58, 0.64) | 0.63 (0.6, 0.66) | 0.02 (0, 0.04) |
| Enterocytes | 0.77 (0.75, 0.78) | 0.75 (0.73, 0.76) | -0.02 (-0.03, -0.01) |
| Endothelial Cells | 0.28 (0.23, 0.33) | 0.29 (0.25, 0.33) | 0.01 (-0.02, 0.04) |

Non-inferiority criteria were set based on inter-pathologist variability for each substance which were then evaluated on 75 μm x 75 μm frames (n = 400 frames). Eosinophil, plasma cell, lymphocyte, enterocyte, and goblet cell performance were considered non-inferior to pathologists if the lower bound of the $\Delta$ F1 confidence interval was $>-0.1$ . Neutrophil and endothelial cell performance were considered non-inferior to pathologists if the lower bound of the $\Delta$ F1 confidence interval was $>-0.2$ . Macrophage, fibroblast and other cell performance were evaluated but considered exploratory substances

**Supplementary Table 4.** IBDExplore mucin model performance in the held-out test set.

|  | **Pathologist vs.**<br>**Pathologist F1 (95% CI)** | **Model vs. Pathologist**<br>**F1 (95% CI)** | **$\Delta F1$ (95% CI)** |
| --- | --- | --- | --- |
| Goblet Cell<br>Mucin | 0.8 (0.79, 0.81) | 0.75 (0.74, 0.76) | -0.05 (-0.05, -0.04) |

Non-inferiority criteria were set based on inter-pathologist variability for each substance which were then evaluated on 400 $\mu\text{m}$ x 400 $\mu\text{m}$ frames (n = 100 frames). Goblet cell mucin performance was considered non-inferior to pathologists if the lower bound of the $\Delta F1$ confidence interval was $>-0.1$ .

**Supplementary Table 5.**
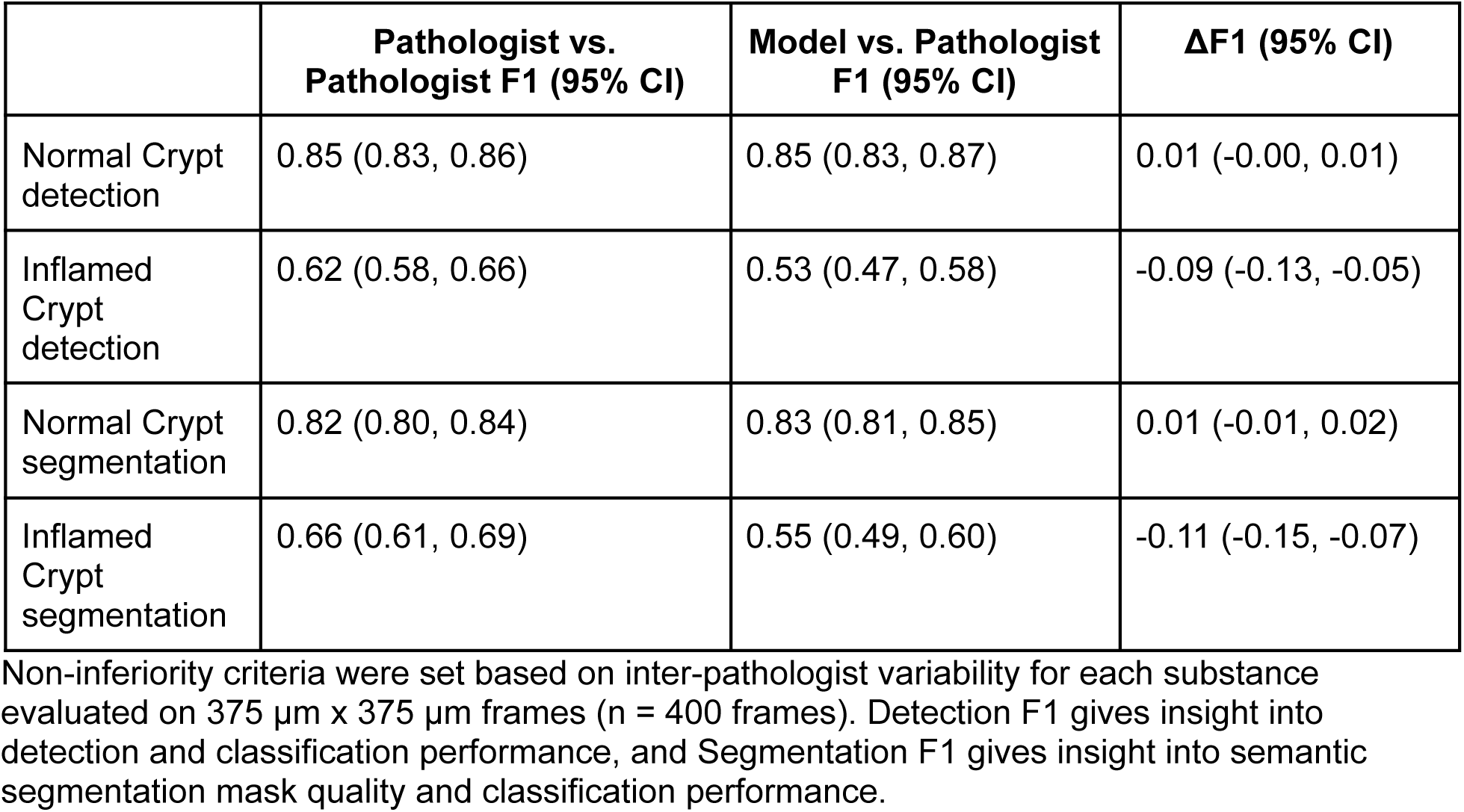
IBDExplore crypt model performance in the held-out test set.

|  | **Pathologist vs.**<br>**Pathologist F1 (95% CI)** | **Model vs. Pathologist**<br>**F1 (95% CI)** | **ΔF1 (95% CI)** |
| --- | --- | --- | --- |
| Normal Crypt<br>detection | 0.85 (0.83, 0.86) | 0.85 (0.83, 0.87) | 0.01 (-0.00, 0.01) |
| Inflamed<br>Crypt<br>detection | 0.62 (0.58, 0.66) | 0.53 (0.47, 0.58) | -0.09 (-0.13, -0.05) |
| Normal Crypt<br>segmentation | 0.82 (0.80, 0.84) | 0.83 (0.81, 0.85) | 0.01 (-0.01, 0.02) |
| Inflamed<br>Crypt<br>segmentation | 0.66 (0.61, 0.69) | 0.55 (0.49, 0.60) | -0.11 (-0.15, -0.07) |

Non-inferiority criteria were set based on inter-pathologist variability for each substance evaluated on 375 μm x 375 μm frames (n = 400 frames). Detection F1 gives insight into detection and classification performance, and Segmentation F1 gives insight into semantic segmentation mask quality and classification performance.

**Supplementary Table 6.**
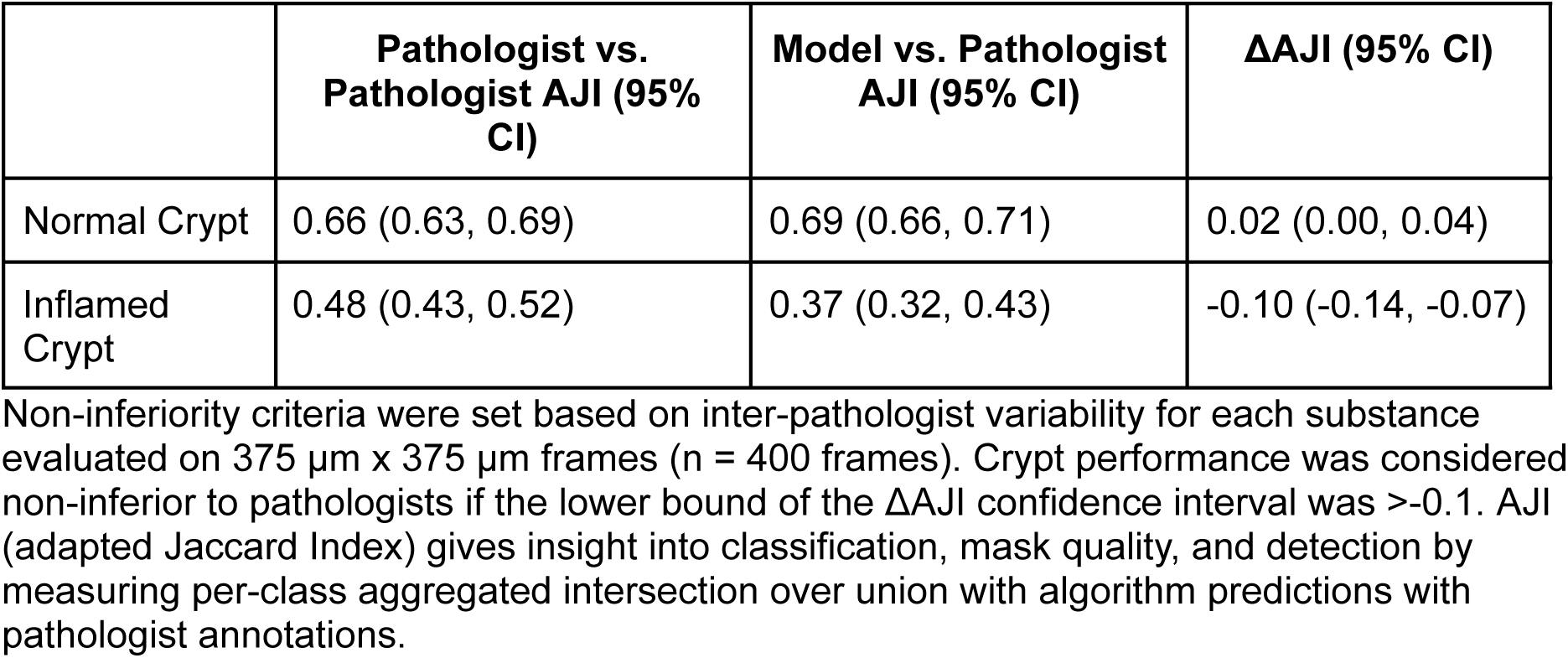
IBDExplore crypt classification model performance in the held-out test set.

**Supplementary Table 7.**
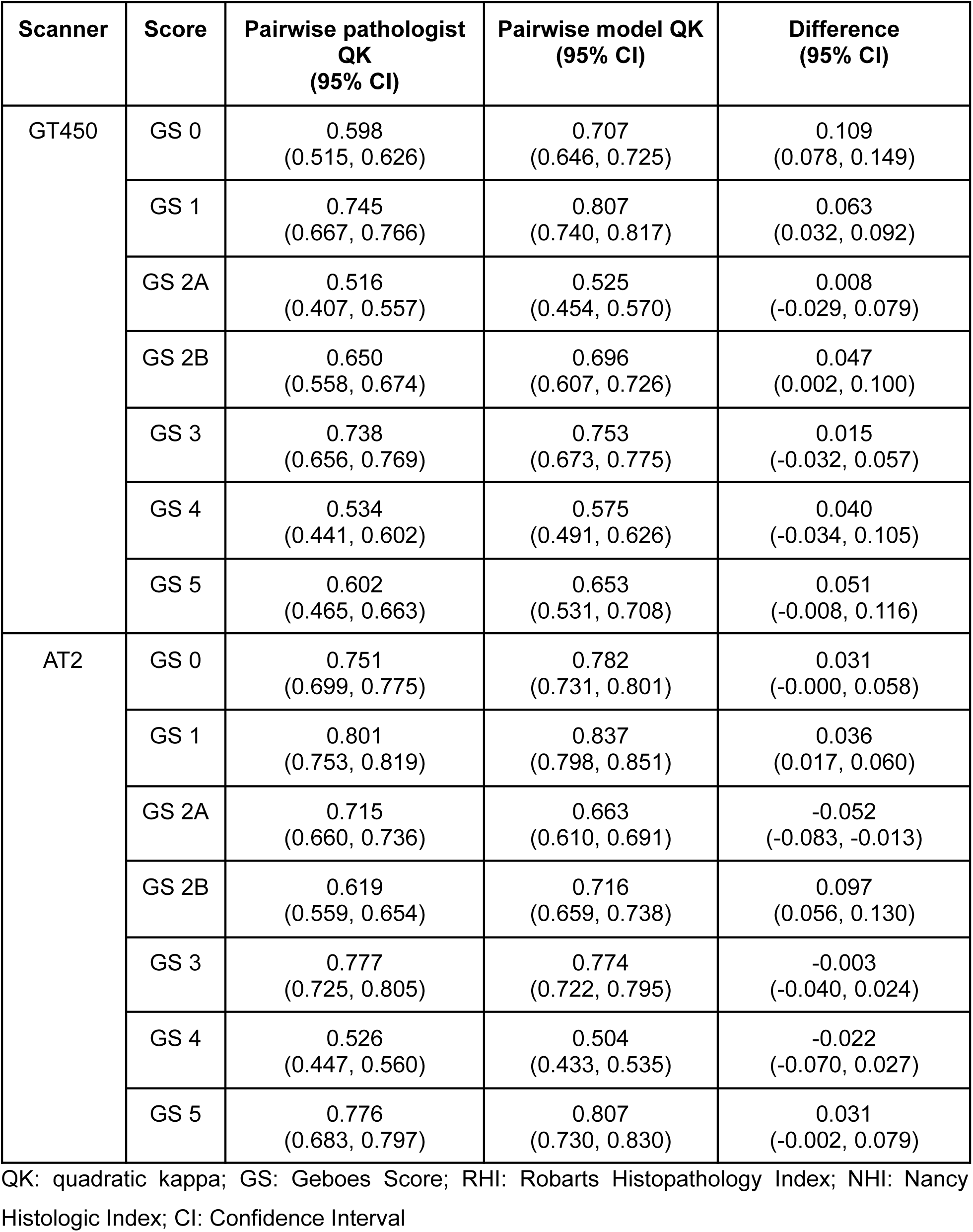
Scanner-specific comparison of manual pathologist Geboes subgrade scores with model predictions in the standalone verification set.

**Supplementary Table 8.** Scanner-specific comparison of manual pathologist grade-level Geboes scores, RHI, and NHI with model predictions with model predictions.

| Scanner | Score | Pairwise pathologist LK (95% CI) | Pairwise model LK (95% CI) | Difference (95% CI) |
| --- | --- | --- | --- | --- |
| GT450 | Overall grade | 0.578<br>(0.502, 0.606) | 0.636<br>(0.560, 0.656) | 0.058<br>(0.019, 0.088) |
|  | RHI | 0.598<br>(0.529, 0.620) | 0.650<br>(0.577, 0.664) | 0.052<br>(0.013, 0.079) |
|  | NHI | 0.649<br>(0.563, 0.676) | 0.657<br>(0.575, 0.683) | 0.008<br>(-0.035, 0.055) |
| AT2 | Overall grade | 0.731<br>(0.673, 0.749) | 0.724<br>(0.667, 0.747) | -0.007<br>(-0.038, 0.029) |
|  | RHI | 0.706<br>(0.660, 0.721) | 0.711<br>(0.670, 0.727) | 0.005<br>(-0.018, 0.031) |
|  | NHI | 0.761<br>(0.706, 0.784) | 0.738<br>(0.682, 0.763) | -0.023<br>(-0.061, 0.013) |

LK: linear kappa; GS: Geboes Score; RHI: Robarts Histopathology Index; NHI: Nancy Histologic Index; CI: Confidence Interval

**Supplemental Table 9.** Scanner-specific comparison of manual pathologist assessment of histologic remission and improvement with model predictions.

| Scanner | Metric |  | Pairwise pathologist (95% CI) | Pairwise model (95% CI) | Difference (95% CI) |
| --- | --- | --- | --- | --- | --- |
| GT450 | 2A Histologic Remission | PPA | 0.743<br>(0.674, 0.802) | 0.845<br>(0.776, 0.898) | 0.102<br>(0.035, 0.159) |
|  |  | NPA | 0.895<br>(0.867, 0.921) | 0.895<br>(0.866, 0.926) | 0.000<br>(-0.021, 0.023) |
|  | 2B Histologic Remission | PPA | 0.843<br>(0.805, 0.883) | 0.860<br>(0.801, 0.900) | 0.017<br>(-0.031, 0.045) |
|  |  | NPA | 0.900<br>(0.855, 0.922) | 0.862<br>(0.807, 0.899) | -0.039<br>(-0.080, 0.006) |
|  | Histologic Improvement | PPA | 0.843<br>(0.798, 0.875) | 0.782<br>(0.727, 0.828) | -0.061<br>(-0.098, -0.021) |
|  |  | NPA | 0.842<br>(0.798, 0.874) | 0.878<br>(0.831, 0.924) | 0.036<br>(-0.002, 0.087) |
| AT2 | 2A Histologic Remission | PPA | 0.863<br>(0.817, 0.895) | 0.874<br>(0.832, 0.918) | 0.011<br>(-0.021, 0.060) |
|  |  | NPA | 0.959<br>(0.945, 0.970) | 0.958<br>(0.940, 0.974) | 0.000<br>(-0.017, 0.015) |
|  | 2B Histologic Remission | PPA | 0.874<br>(0.836, 0.904) | 0.879<br>(0.836, 0.930) | 0.006<br>(-0.034, 0.055) |
|  |  | NPA | 0.936<br>(0.918, 0.952) | 0.893<br>(0.863, 0.921) | -0.042<br>(-0.067, -0.019) |
|  | Histologic Improvement | PPA | 0.879<br>(0.849, 0.906) | 0.848<br>(0.802, 0.888) | -0.032<br>(-0.069, 0.006) |
|  |  | NPA | 0.917<br>(0.897, 0.935) | 0.910<br>(0.883, 0.937) | -0.006<br>(-0.031, 0.019) |

PPA: positive percent accuracy; NPA: negative percent accuracy; CI: confidence interval

**Supplemental Table 10.** IBDExplore HIFs selected for comparison to Geboes subgrades.

| **Geboes subgrade** | **Histology measured** | **IBDExplore HIF** |
| --- | --- | --- |
| GS 0 | Crypt architectural changes | • Standard deviation in crypt area<br>• Mean crypt solidity |
| GS 1 | Chronic inflammatory infiltrate | • Fraction of total cells in mucosa predicted to be lymphocytes<br>• Fraction of total cells in mucosa predicted to be plasma cells |
| GS 2A | Eosinophils in lamina propria | • Fraction of total cells in lamina propria predicted to be eosinophils |
| GS 2B | Neutrophils in lamina propria | • Fraction of total cells in lamina propria predicted to be neutrophils |
| GS 3 | Neutrophils in epithelium | • Fraction of total crypt cells predicted to be neutrophils<br>• Fraction of total crypts predicted to be inflamed |
| GS 4 | Crypt destruction | • Fraction of mucosal area predicted to be crypt abscess<br>• Proportion of crypt area predicted to be infiltrated |
| GS 5 | Erosions and ulcerations | • Relative area of complete granulation tissue, erosion, and ulceration in lamina propria |

## Supplementary Figures

**Supplementary Figure 1.**
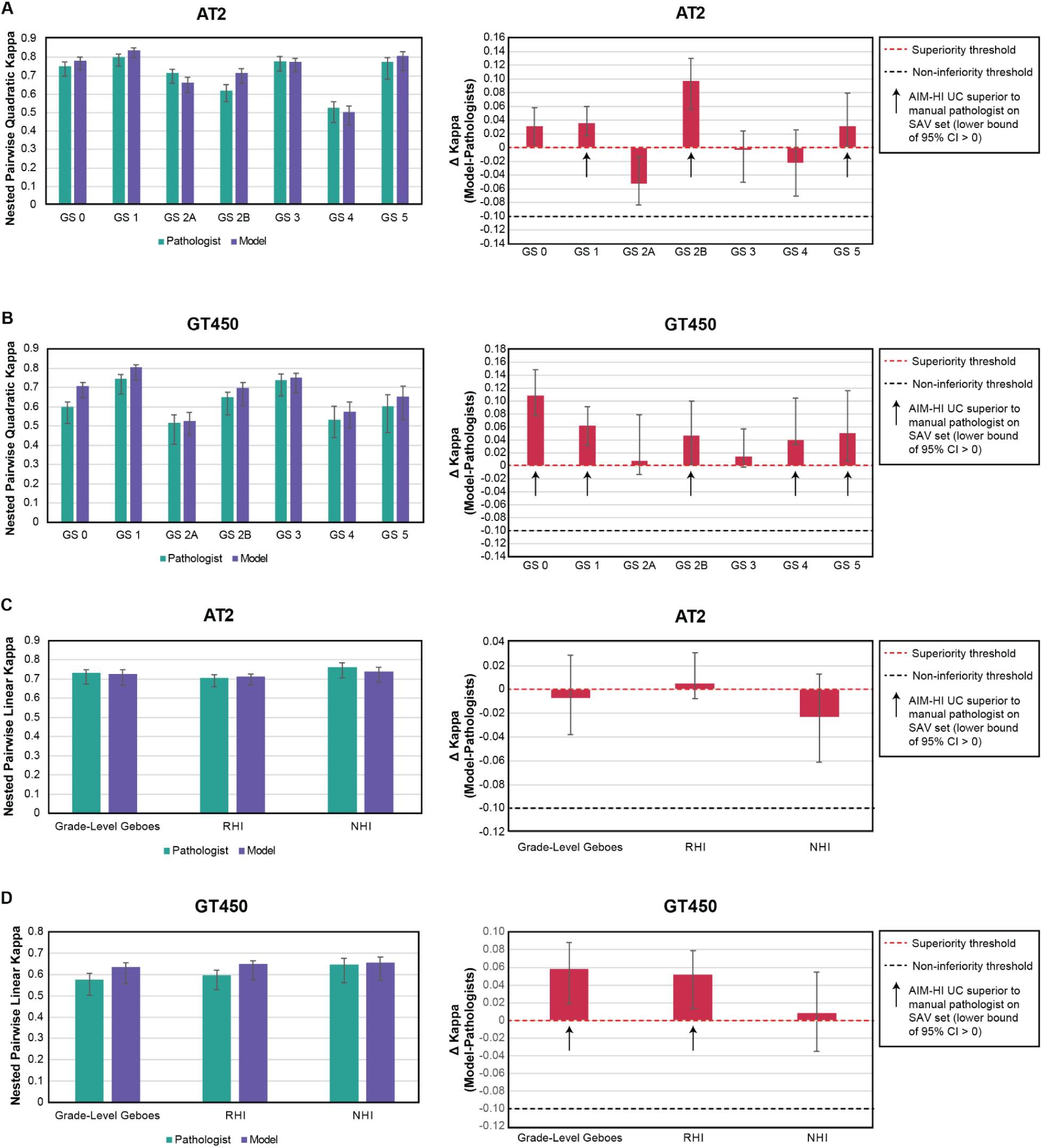
AIM-HI UC model performance compared to pathologists for histologic scoring on AT2 and GT450 scanners. A) Comparison of nested pairwise quadratic kappa values from pathologist manual scoring and AIM-HI UC predictions of Geboes subgrades on AT2-scanned slides. Difference in quadratic kappa between model and pathologist for Geboes subgrade determination on AT2-scanned slides is shown. B) Comparison of nested pairwise linear kappa values from pathologist manual scoring and AIM-HI UC predictions of overall grade-level Geboes score, RHI, and NHI on GT450 scanned slides. Difference in linear kappa between model and pathologist for overall grade-level Geboes score, RHI, and NHI determination on AT2-scanned slides is shown. C) Comparison of nested pairwise quadratic kappa values from pathologist manual scoring and AIM-HI UC predictions of Geboes subgrades on AT2-scanned slides. Difference in quadratic kappa between model and pathologist for Geboes subgrade determination on AT2-scanned slides is shown. D) Comparison of nested pairwise linear kappa values from pathologist manual scoring and AIM-HI UC predictions of overall grade-level Geboes score, RHI, and NHI on GT450 scanned slides. Difference in linear kappa between model and pathologist for overall grade-level Geboes score, RHI, and NHI determination on AT2-scanned slides is shown. Error bars represent 95% confidence intervals. GS: Geboes score, RHI: Robarts Histopathology Index, NHI: Nancy Histology Index, CI: confidence interval.

**Supplementary Figure 2.**
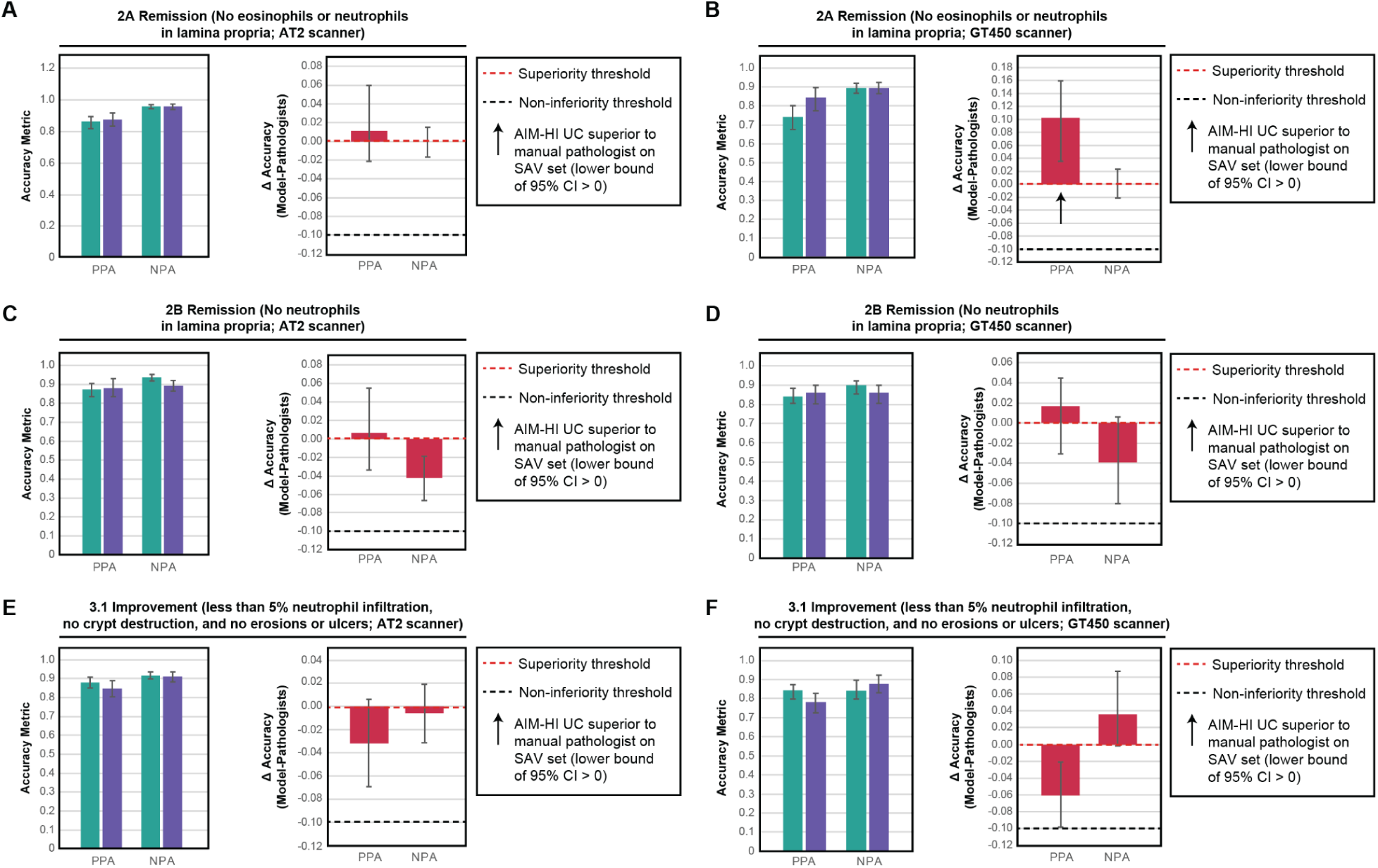
AIM-HI UC model performance compared to pathologists for determination of histologic remission status on AT2 and GT450 scanners. A) Comparison of positive percent agreement (PPA) and negative percent agreement (NPA) from pathologist manual scoring and AIM-HI UC prediction of 2A histologic remission on AT2-scanned slides. Difference in PPA and NPA between model and pathologist for 2A histologic remission on AT2-scanned slides is shown. B) Comparison of PPA and NPA from pathologist manual scoring and AIM-HI UC prediction of 2A histologic remission on GT450-scanned slides. Difference in PPA and NPA between model and pathologist for 2A histologic remission on GT450-scanned slides is shown. C) Comparison of PPA and NPA from pathologist manual scoring and AIM-HI UC prediction of 2B histologic remission on AT2-scanned slides. Difference in PPA and NPA between model and pathologist for 2B histologic remission on AT2-scanned slides is shown. D) Comparison of PPA and NPA from pathologist manual scoring and AIM-HI UC prediction of 2B histologic remission on GT450-scanned slides. Difference in PPA and NPA between model and pathologist for 2B histologic remission on GT450-scanned slides is shown. E) Comparison of PPA and NPA from pathologist manual scoring and AIM-HI UC prediction of histologic improvement on AT2-scanned slides. Difference in PPA and NPA between model and pathologist for histologic improvement on AT2-scanned slides is shown. F) Comparison of PPA and NPA from pathologist manual scoring and AIM-HI UC prediction of histologic improvement on GT450-scanned slides. Difference in PPA and NPA between model and pathologist for histologic improvement on GT450-scanned slides is shown. Error bars represent 95% confidence intervals. PPA: positive percent agreement, NPA: negative percent agreement; CI: confidence interval.

**Supplementary Figure 3.**
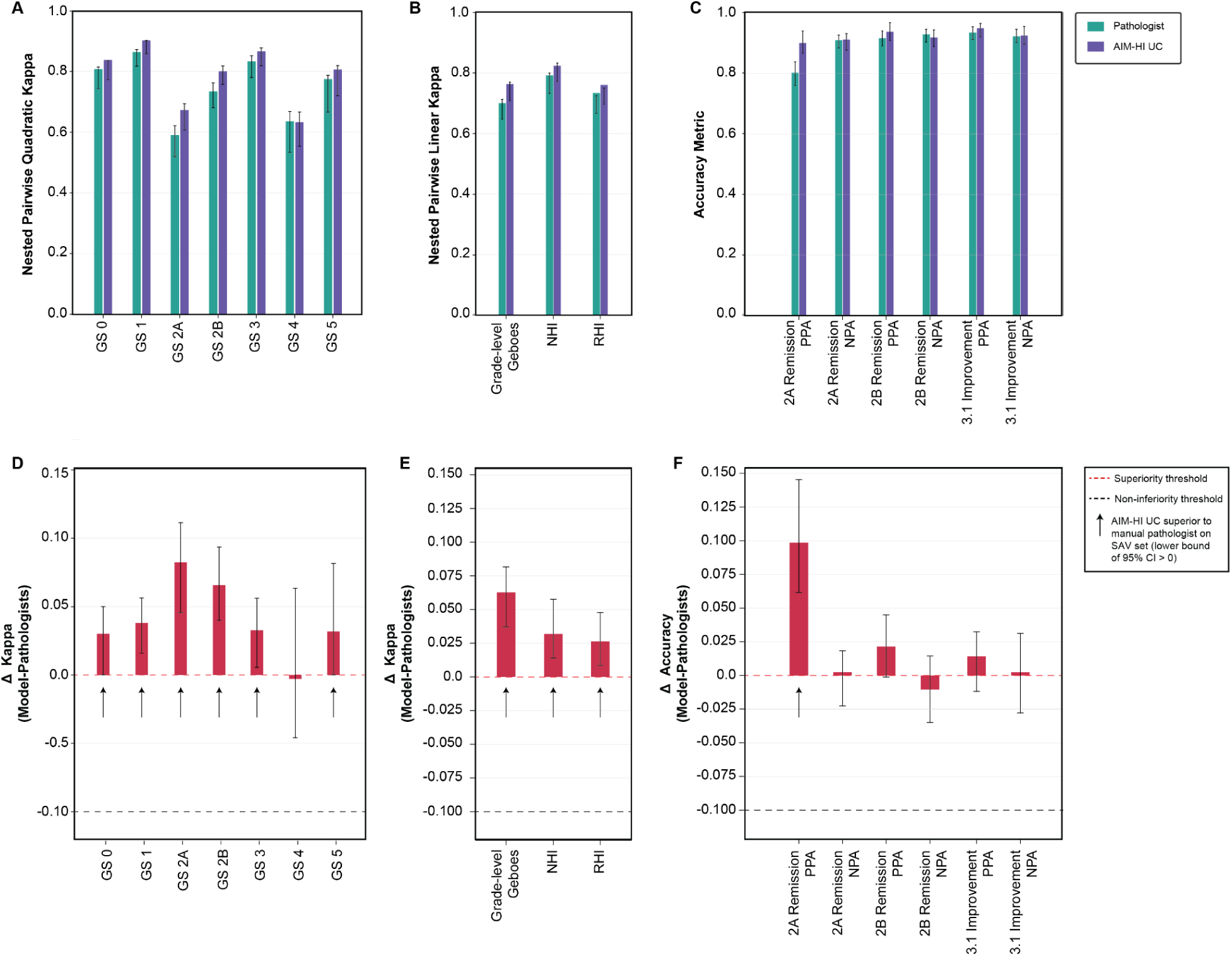
AIM-HI UC model performance compared to pathologists for histologic scoring in the held-out test set (n = 578). A) Comparison of nested pairwise quadratic kappa values from pathologist manual scoring and AIM-HI UC predictions of Geboes subgrades. B) Comparison of nested pairwise linear kappa values from pathologist manual scoring and AIM-HI UC predictions of overall grade-level Geboes score, RHI, and NHI. C) Comparison of PPA and NPA from pathologist manual scoring and AIM-HI UC prediction of 2A histologic remission, 2B histologic remission, and 3.1 histologic improvement. D) Difference in quadratic kappa between model and pathologist for Geboes subgrade determination. E) Difference in linear kappa between model and pathologist for overall grade-level Geboes score, RHI, and NHI determination. F) Difference in quadratic kappa between model and pathologist for 2A histologic remission, 2B histologic remission, and 3.1 histologic improvement. Error bars represent 95% confidence intervals. GS: Geboes Subgrade; PPA: positive percent agreement; NPA: negative percent agreement.

**Supplementary Figure 4.**
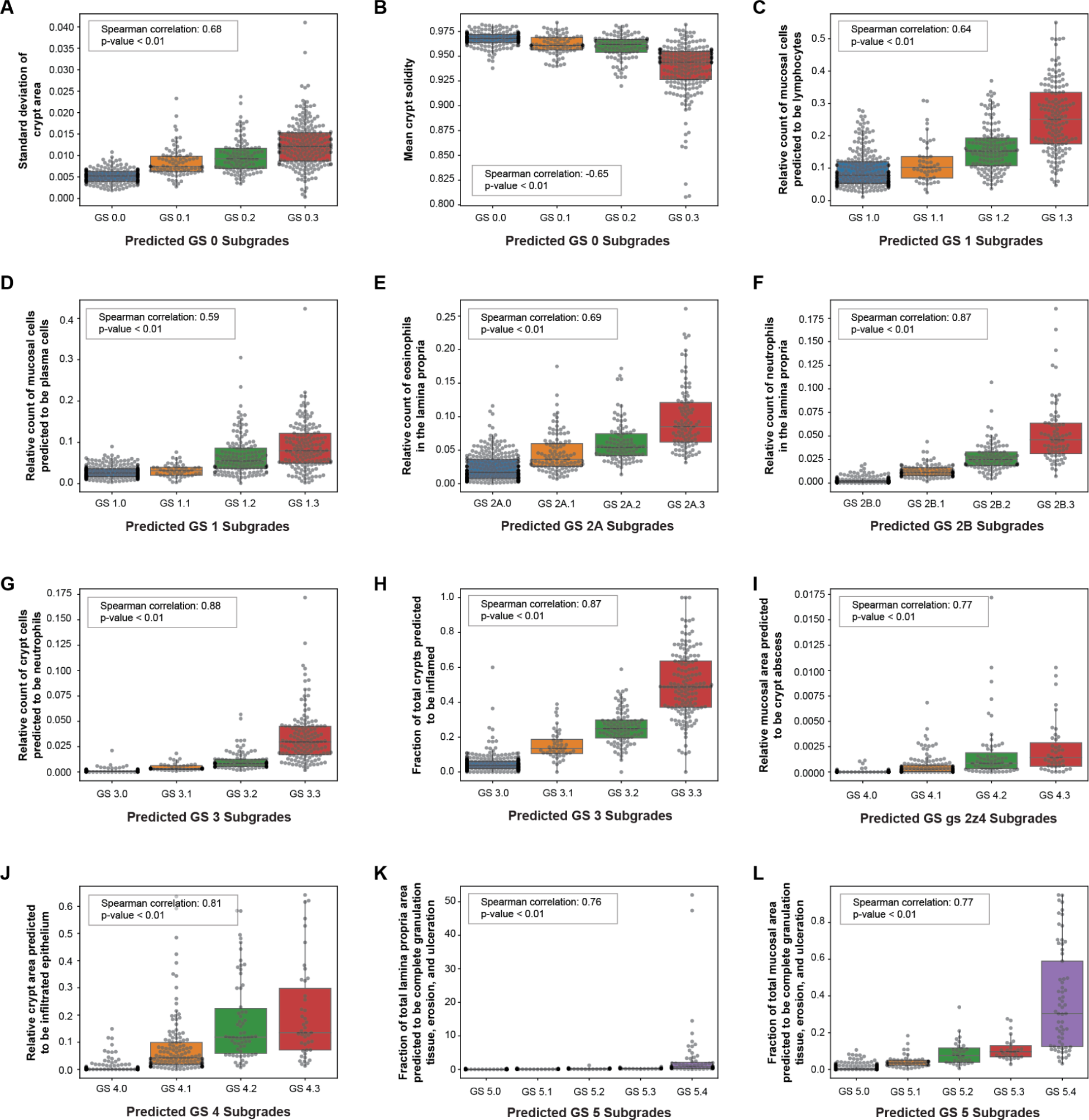
Correlation between AIM-HI UC-derived Geboes subgrade subscores and AI-quantified features of the inflammatory microenvironment. A-B) Correlation between AIM-HI UC-predicted GS 0 subscores and IBDExplore-quantified A) standard deviation of crypt area and B) mean crypt solidity. C-D) Correlation between AIM-HI UC-predicted GS 1 subscores and IBDExplore-quantified C) relative count of mucosal cells predicted to be lymphocytes and D) relative count of mucosal cells predicted to be plasma cells. E) Correlation between AIM-HI UC-predicted GS 2A subscores and IBDExplore-quantified relative count of eosinophils in the lamina propria. F) Correlation between AIM-HI UC-predicted GS 2B subscores and IBDExplore-quantified relative count of neutrophils in the lamina propria. G-H) Correlation between AIM-HI UC-predicted GS 3 subscores and IBDExplore-quantified G) relative count of crypt cells predicted to be neutrophils and H) fraction of total crypts predicted to be inflamed. I-J) Correlation between AIM-HI UC-predicted GS 4 subscores and IBDExplore-quantified I) relative mucosal area predicted to be crypt abscess and J) relative crypt area predicted to be infiltrated epithelium. K-L) Correlation between AIM-HI UC-predicted GS 5 subscores and IBDExplore-quantified K) fraction of total lamina propria area predicted to be complete granulation tissue, erosion, and ulceration and L) fraction of total mucosal area predicted to be complete granulation tissue, erosion, and ulceration. Error bars represent 95% confidence intervals. GS: Geboes Score.

**Supplementary Figure 5.**
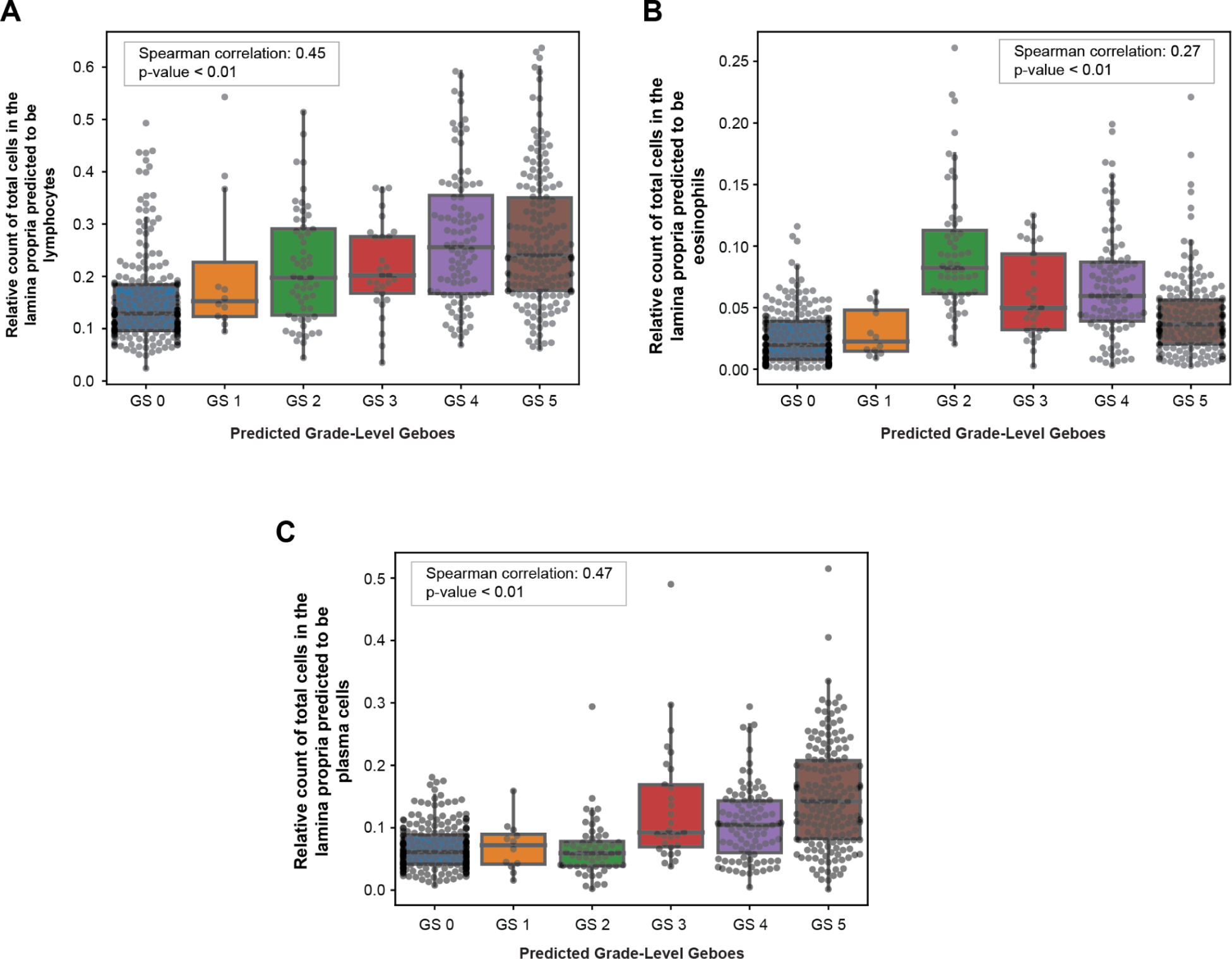
Correlation between AIM-HI UC grade level Geboes scores and AI-quantified cell types in the lamina propria. A) Relative count of lymphocytes localized within the lamina propria, B) relative count of eosinophils localized within the lamina propria, and C) relative count of plasma cells localized within the lamina propria. Error bars represent 95% confidence intervals. GS: Geboes Score.

